# Anaesthetic managment and clinical outcomes of parturients with COVID-19: a multicentre, retrospective, propensity score matched cohort study

**DOI:** 10.1101/2020.03.24.20042176

**Authors:** Yuan Zhang, Rong Chen, Jie Wang, Yuan Gong, Qin Zhou, Hui-hui Cheng, Zhong-yuan Xia, Xiangdong Chen, Qing-tao Meng, Daqing Ma

**Affiliations:** Department of Anaesthesiology, Renmin Hospital of Wuhan University, Wuhan 430060, China; Department of Anaesthesiology, East Hospital, Renmin Hospital of Wuhan University, Wuhan 430060, China; Department of Anaesthesiology, Union Hospital, Tongji Medical College, Huazhong University of Science and Technology, Wuhan 430056, China; Department of Anaesthesiology, Yichang Central People’s Hospital, The First College of Clinical Medical Science, China Three Gorges University, Yichang 443003, China; Division Anaesthetics, Pain Medicine and Intensive Care, Department of Surgery and Cancer, Faculty of Medicine, Imperial College London, Chelsea & Westminster Hospital, London, United Kingdom

**Author notes:** Corresponding Author: Dr. Qing-tao Meng, Department of Anaesthesiology, Renmin Hospital of Wuhan University, Wuhan 430060, PR. China. These authors contributed equally to this work (Yuan Zhang, Rong Chen, Jie Wang, Yuan Gong). Qing-tao Meng, Xiangdong Chen and Daqing Ma share senior authorship.

**Keywords:** COVID-19, Cesarean delivery, Anaesthesia, Propensity score matching, Cohort study

## Abstract

**Objective:** To analyse the clinical features of COVID-19 parturients, and to compare anaesthetic regimen and clinical outcomes in parturients with or without COVID-19 undergoing cesarean delivery.

**Methods:** Data were extracted from the electronic medical record of 3 medical institutions in Hubei Province, China, from June 1, 2019 to March 20, 2020 according to inclusion and exclusion criteria. After propensity score matching with demographics, the clinical and laboratory characteristics of parturients with or without COVID-19 were analysed. The anaesthetic regimen and clinical outcomes of themselves and their infants were compared in these two groups of parturients.

**Results:** A total of 1,588 patients without SARS-CoV-2 infection undergoing cesarean delivery were retrospectively included. After achieving a balanced cohort through propensity score matching, 89 patients (COVID-19 group), who were diagnosed with COVID-19 by SARS-CoV-2 nucleic acid test and CT scan matched with 173 patients without COVID-19 (Control group). The SARS-CoV-2 infected parturients in the early stages of COVID-19 outbreak was much more than during the later stage. The main clinical characteristics of parturients with COVID-19 were fever (34.8%), cough (33.7%), an increased plasma CRP (52.8%) and a decreased lymphocyte counting (33.7%). A high rate of emergency and a high incidence of anaesthesia-related complications, such as pharyngalgia, multiple puncture, intraoperative hypotension, nausea, vomiting, vertigo and chills in the COVID-19 parturients. In addition, the parturients with COVID-19 had a long duration of operation and hospital stay, and an increased intraoperative oxytocin utilization and postoperative oxygen therapy. The newborns from the SARS-CoV-2 infected mothers, who received general anaesthesia, had a high risk of Apgar score ≤ 8 at 1 and 5 minutes after delivery and a higher rate of neonatal intensive care unit (NICU) admission.

**Conclusions:** Anaesthesia-related complications occur more frequently in the COVID-19 parturients and their newborns have a high risk of distress.

## Introduction

Since December 2019, a novel coronavirus (SARS-CoV-2) infection disease (COVID-19) has been reported with an outbreak in Wuhan and now has become global pandemic. More than 50 countries have declared a state of emergency, and some countries have even declared a state of “war”. As of March 21, 2020, a total 266,073 patients of COVID-19 have been diagnosed in the world^1^. The clinical manifestations of SARS-Cov-2 infection range from asymptomatic (no symptoms) mild to severe organ dysfunction such as acute respiratory distress syndrome (ARDS) that can lead to death^2–4^. The common symptoms including fever, dry cough, dyspnea, fatigue, myalgia, and lymphopenia in infected patients. The typical abnormality of CT scan was bilateral distribution of patchy shadows and ground-glass opacity^5^. A study has reported that the pathological features of COVID-19 greatly are similar with those seen in SARS and Middle Eastern respiratory syndrome (MERS) coronavirus infection, such as the bilateral diffuse alveolar damage with cellular fibromyxoid exudates, pulmonary oedema with hyaline membrane formation^6^.

As a member of the coronavirus family, SARS-CoV-2 is now spread human-to-human with higher infectivity than MERS and SARS but a lower fatality rate^7^. COVID-19 is highly transmissible disease via respiratory droplets, aerosols and close contact. Parturients were considered to be susceptible of SARS-CoV-2 virus and COVID-19 can threaten to the health of maternal and infant. Although the clinical characteristics of COVID-19 in pregnant women are similar with non-pregnant females, COVID-19 resulted in additional challenges for obstetric anaesthesia under cesarean delivery as reported in our previous study^8^. However, the comparison between parturients undergoing cesarean delivery with versus without COVID-19 are still undefined and the clinical features of parturients with COVID-19 require further investigation. In present study, we retrospectively analyzed and compared the anaesthetic data and outcomes for parturients with or without COVID-19 in multicentre during and after cesarean delivery.

## Methods

### Patients

A multicentre, retrospective, propensity score matched cohort study was conducted to compare the anaesthetic practices in patients with versus without COVID-19 undergoing cesarean delivery. Data were obtained from the electronic medical records of Renmin hospital of Wuhan university, Union hospital affiliated to Tongji medical college of Huazhong university of science and technology and Yichang central people’s hospital, Yichang, Hubei. The electronic search was supplemented by manual chart review when needed. The exclusion criteria included preoperative coexistent diseases, pregnancy complications, stillbirth and fetal congenital abnormality. The patients’ data were harvested from June 1, 2019 to March 20, 2020. The study was approved by the Institutional Review Board at Renmin hospital of Wuhan University (No. WDRY2020-K077) and received an informed consent exemption. All clinical data were independently collected and crossed check by two investigators in each institution.

### Data collection

The data of maternal demographics, clinical characteristics and anaesthesia management, were collected. Maternal and infant clinical outcomes were also recorded.

### Propensity score matching

A propensity score matching method was adopted to screen the counterparts of cases in parturients without COVID-19 using the nearest neighbor matching (caliper 0.2). The matching ratio was 1: 2. The matching factors included maternal age, body mass index (BMI), gestational age, times of cesarean delivery, the number of gestations and ASA classification.

### Statistical analysis

All categorical variables were expressed as number, and all quantitative data were expressed as mean ± standard deviation. Propensity score matching was implemented with Stata 15. Then the matched data were statistically analyzed using Graphpad 8.0.1 (GraphPad Software, San Diego, USA). The two-tailed *t* test was done for continuous variables and Chi-squared test (or Fisher’s exact test) for categorical variables. A *p* value less than 0.05 was considered to be statistically significant.

## Results

### The clinical characteristics of COVID-19 group

A total of 1,588 patients undergoing cesarean delivery were retrospectively included in the study. Firstly, the number of parturients with COVID-19 by the time segment was calculated (**Figure 1**). The COVID-19 cases were gradually decreased after reaching the peak (39 cases) in the first half of February 2020. In the March 2020, there were only 9 COVID-19 cases underwent cesarean delivery. A total of 89 parturients was diagnosed with COVID-19 by SARS-CoV-2 nucleic acid test and CT imaging. However, only 21(23.6%) cases had a history of epidemic exposure to SARS-CoV-2. There are 31(34.8%) cases had persistent fever and 30(33.7%) cases had intermittent cough. The rate of other symptoms, such as fatigue, chest distress, dyspnoea and diarrhea were less than 10%. The most common changes in the laboratory test was an increased change of plasma CRP (52.8%) followed by a decreased counting of lymphocyte (33.7%) and leukocytosis (23.6%). In addition, some cases had mild liver and kidney dysfunction (**Table 1**).

**Table 1.** The clinical characteristics of COVID-19 group (n=133)

|  | COVID-19 group |  | Total |
| --- | --- | --- | --- |
|  | GA (n=11) | IA (n=78) | n(%) |
| Epidemiological exposure to SARS-CoV-2, n(%) | 2(18.2) | 19(24.4) | 21(23.6) |
| Signs and symptoms |  |  |  |
| Fever, n(%) | 1(9.1) | 30(38.5) | 31(34.8) |
| Cough, n(%) | 8(72.7) | 22(28.2) | 30(33.7) |
| Fatigue, n(%) | 2(18.2) | 6(7.7) | 8(9.0) |
| Chest distress, n(%) | 6(54.5) | 6(7.7) | 12(13.5) |
| Dyspnoea, n(%) | 3(27.3) | 6(7.7) | 9(10.1) |
| Diarrhea, n(%) | 3(27.3) | 4(5.1) | 7(7.9) |
| Laboratory characteristics |  |  |  |
| Leukocytosis > 10×10<sup>9</sup>/L, n(%) | 11(100) | 10(12.8) | 21 (23. 6) |
| Lymphocyte percentage < 20%, n(%) | 7(63.6) | 23(29.5) | 30 (33. 7) |
| CRP concentrations > 10 mg/L, n(%) | 9(81.8) | 38(43.6) | 47(52.8) |
| ALT > 40U/L, n(%) | 1(9.1) | 3(3.8) | 4 (4. 5) |
| AST > 40U/L, n(%) | 0 | 5(6.4) | 5(5.6) |
| BUN > 8.3mM, n(%) | 0 | 2(2.6) | 2(2.2) |
| SCr > 84μM, n(%) | 0 | 3(3.8) | 3(3.4) |
| CT evidence of pneumonia, n(%) | 11(100) | 78(100) | 89(100) |
| Positive throat swab of SARS-CoV-2 nucleic acid test | 11(100) | 78(100) | 89(100) |

GA: general anaesthesia, IA: intrathecal anaesthesia

**Figure 1.**
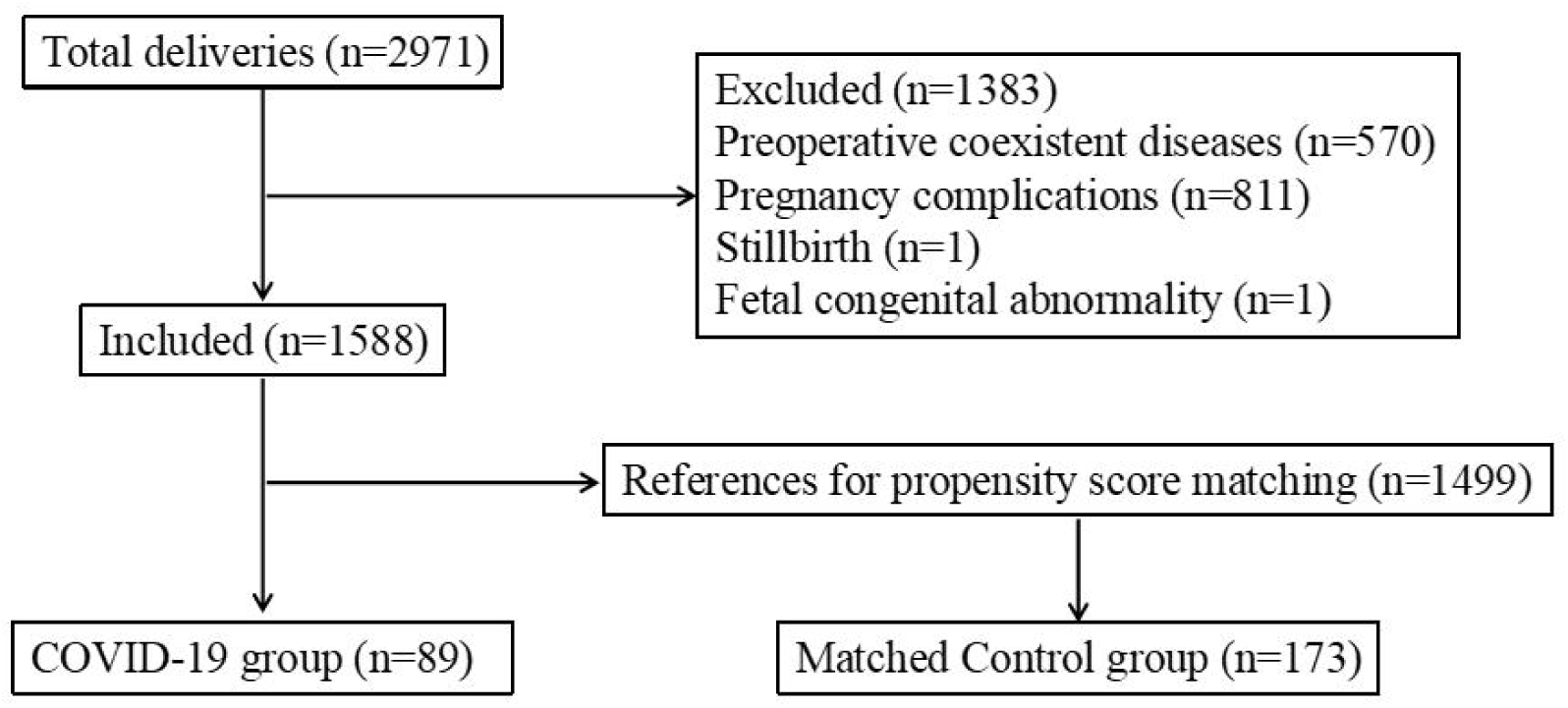
The flowchart of data extraction.

### Propensity matching scores

A total of 1,588 cases without COVID-19 were included as references for propensity score matching. After achieving a balanced cohort through propensity score matching, 89 patients with COVID-19 (the COVID-19 group) and 173 patients without COVID-19 (the Control group) were obtained. Among them, 5 patients with COVID-19 successfully matched only one counterpart because of the strict matching conditions and others successfully matched two counterparts. A flowchart was shown in **Figure 2**. The distributions of patient characteristics before and after propensity score matching are shown in **Table 2**. After, propensity score matching, the baseline characteristics between the two groups were comparable (all *p*>0.05).

**Table 2.** The baseline characteristics of two groups before and after matching

|  | COVID-19 group |  | Unmatched control group |  | *P* |  | Matched control group |  | *P* |  |
| --- | --- | --- | --- | --- | --- | --- | --- | --- | --- | --- |
|  | GA (n=11) | IA (n=78) | GA (n=53) | IA (n=1446) | GA | IA | GA (n=18) | IA (n=155) | GA | IA |
| Age (years), mean(SD) | 33.70(3.22) | 30.05(4.4) | 32.53(2.19) | 29.6(2.12) | 0.144 | 0.092 | 33.54(3.67) | 29.98(3.02) | 0.906 | 0.887 |
| BMI (kg/m2), mean(SD) | 24.58(4.15) | 23.21(3.87) | 23.06(3.42) | 24.62(2.02) | 0.208 | <0.05 | 24.50(3.11) | 23.01(3.09) | 0.953 | 0.669 |
| Gestational age (weeks), mean(SD) | 38.09(1.21) | 38.54(1.60) | 39.51(2.08) | 38.92(1.44) | 0.033 | 0.024 | 38.24(1.79) | 38.62(0.94) | 0.808 | 0.632 |
| Times of cesarean delivery |  |  |  |  |  |  |  |  |  |  |
| Primary, n(%) | 2(18.2) | 49(62.8) | 14(26.4) | 642(44.4) | 0.716 | 0.002 | 3(16.7) | 98(63.2) | >0.99 | >0.99 |
| Repeated, n(%) | 9(81.8) | 29(37.2) | 39(73.6) | 804(55.6) |  |  | 15(83.3) | 57(36.8) |  |  |
| Number of gestations |  |  |  |  |  |  |  |  |  |  |
| Singleton, n(%) | 11(100) | 77(98.7) | 51(96.2) | 1407(97.3) | >0.99 | 0.719 | 18(100) | 154(99.4) | >0.99 | >0.99 |
| Twin, n(%) | 0 | 1(1.3) | 2(3.8) | 39(2.7) |  |  | 0 | 1(0.6) |  |  |
| ASA classification |  |  |  |  |  |  |  |  |  |  |
| I, n(%) | 4(36.4) | 14(17.9) | 24(45.3) | 952(65.8) | 0.743 | <0.05 | 6(33.3) | 30(19.4) | >0.99 | 0.861 |
| II, n(%) | 7(63.6) | 64(82.1) | 29(54.7) | 494(34.2) |  |  | 12(66.7) | 125(80.6) |  |  |
| III, n(%) | 0 | 0 |  | 0 |  |  | 0 | 0 |  |  |
| IV, n(%) | 0 | 0 |  | 0 |  |  | 0 | 0 |  |  |
| V, n(%) | 0 | 0 |  | 0 |  |  | 0 | 0 |  |  |

GA: general anaesthesia, IA: intrathecal anaesthesia

**Figure 2.**
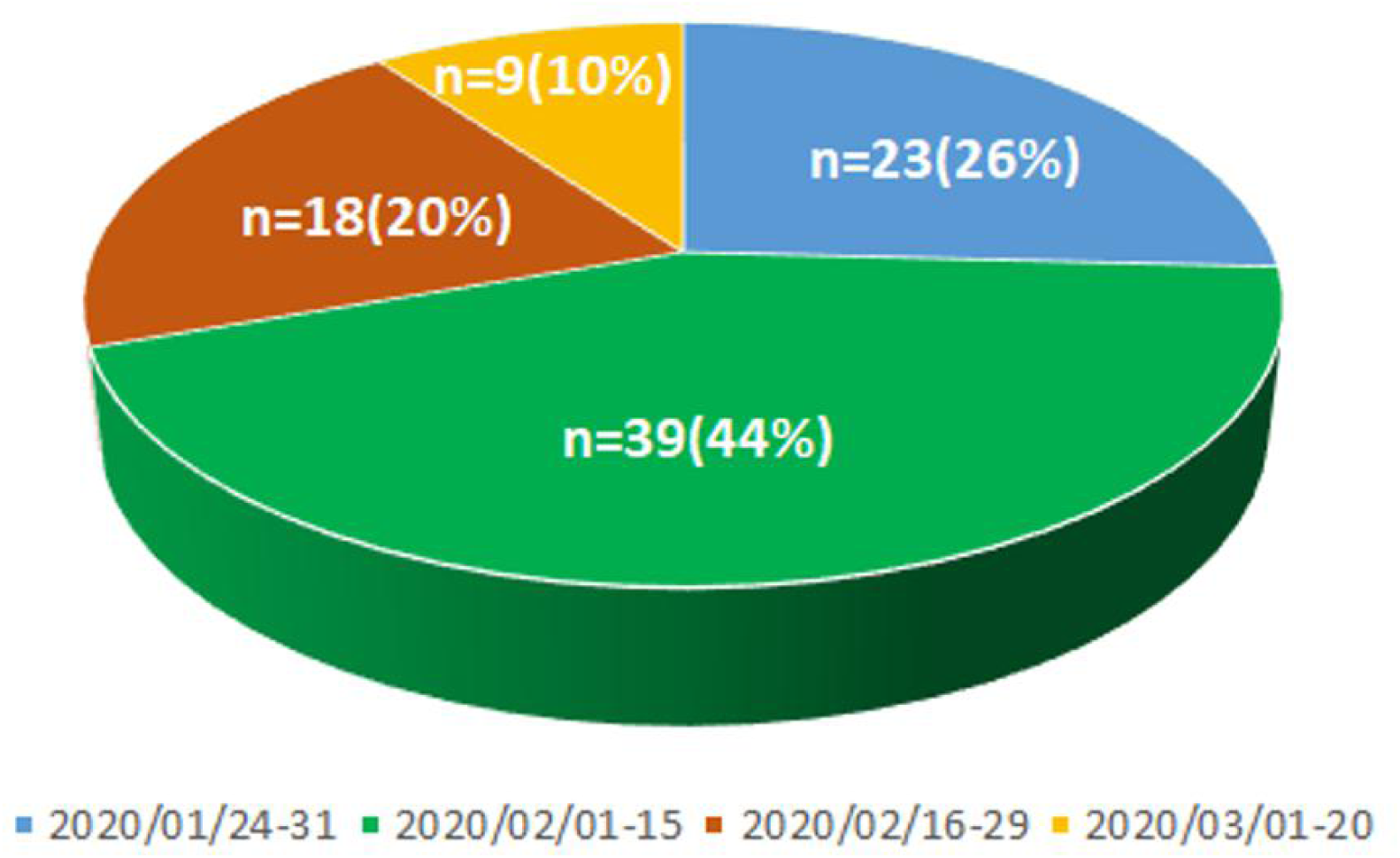
The occurrence of parturients with COVID-19 with the time segment during the peak outbreak.

### Anaesthetic techniques

There were 28(31.5%) cases received elective cesarean delivery, and 61(68.5%) cases underwent emergency cesarean delivery of the COVID-19 parturients. Among of them, 8(9.0%) cases received spinal anaesthesia, 20(22.5%) cases received epidural anaesthesia, 11(12.4%) cases received general anaesthesia induced by intravenous anaesthetic (5.6%) and inhalational anaesthesia (6.7%). In the Control group, 108(62.5%) cases underwent elective cesarean delivery and 63(37.6%) cases received emergency cesarean delivery. Of those, 38(22.0%) cases received spinal anaesthesia, 70(40.5%) cases received epidural anaesthesia, 18(10.4%) cases received general anaesthesia induced by intravenous induction (4.6%) and inhalation induction (5.8%). No significant differences of anaesthetic tecniques were found between two groups. However, the rate of emergency anaesthesia was significantly increased in the COVID-19 parturients (*p*<0.05) (**Table 3**).

**Table 3.** The comparison of anaesthetic mode between two groups after matching

|  | COVID-19 group (n=89) | Control group (n=173) | *P* |
| --- | --- | --- | --- |
| Elective cesarean delivery, n(%) | 28(31.5) | 108(62.5) | 0.679<sup>a</sup> |
| Intravenous induction, n(%) | 0 | 0 |  |
| Inhalational induction, n(%) | 0 | 0 |  |
| Spinal anaesthesia, n(%) | 8(9.0) | 38(22.0) | 0.655 |
| Epidural anaesthesia, n(%) | 20(22.5) | 70(40.5) |  |
| Emergency cesarean delivery, n(%) | 61(68.5) | 65(37.6) | <0.05<sup>b</sup> |
| Intravenous induction, n(%) | 5(5.6) | 8(4.6) | >0.99 |
| Inhalational induction, n(%) | 6(6.7) | 10(5.8) |  |
| Spinal anaesthesia, n(%) | 26(29.2) | 20(11.6) | 0.412 |
| Epidural anaesthesia, n(%) | 24(27.0) | 27(15.6) |  |

<sup>a</sup> the rate of general anaesthesia versus intrathecal anaesthesia between two groups after matching
<sup>b</sup> the rate of elective versus emergency cesarean delivery between two groups after matching

### Anaesthetic management

The length of intrathecal anaesthetic procedure of COVID-19 group was significant longer than the Control group (*p*<0.05). The anaesthetic procedure-related complications, including pharyngalgia and multiple punctures, were significantly increased in the COVID-19 group when compared with the Control group (both *p*<0.05). There was no significant difference in the incidence of unintentional dural puncture, catheter misplacement and nerve root irritation between the two groups. Among the cases who underwent intrathecal anaesthetic procedure, the incidence of adverse responses, including hypotension, nausea, vertigo, chills and vomiting were significantly higher in the COVID-19 group than that in the Control group (all *p*<0.05). However, the incidence of hypotension was only significantly increased in the COVID-19 patients who receiving general anaesthesia, when compared with the Controls. None other complications, e.g. hypoxia and serious arrhythmia, were noticed during operation (**Table 4)**.

**Table 4.**
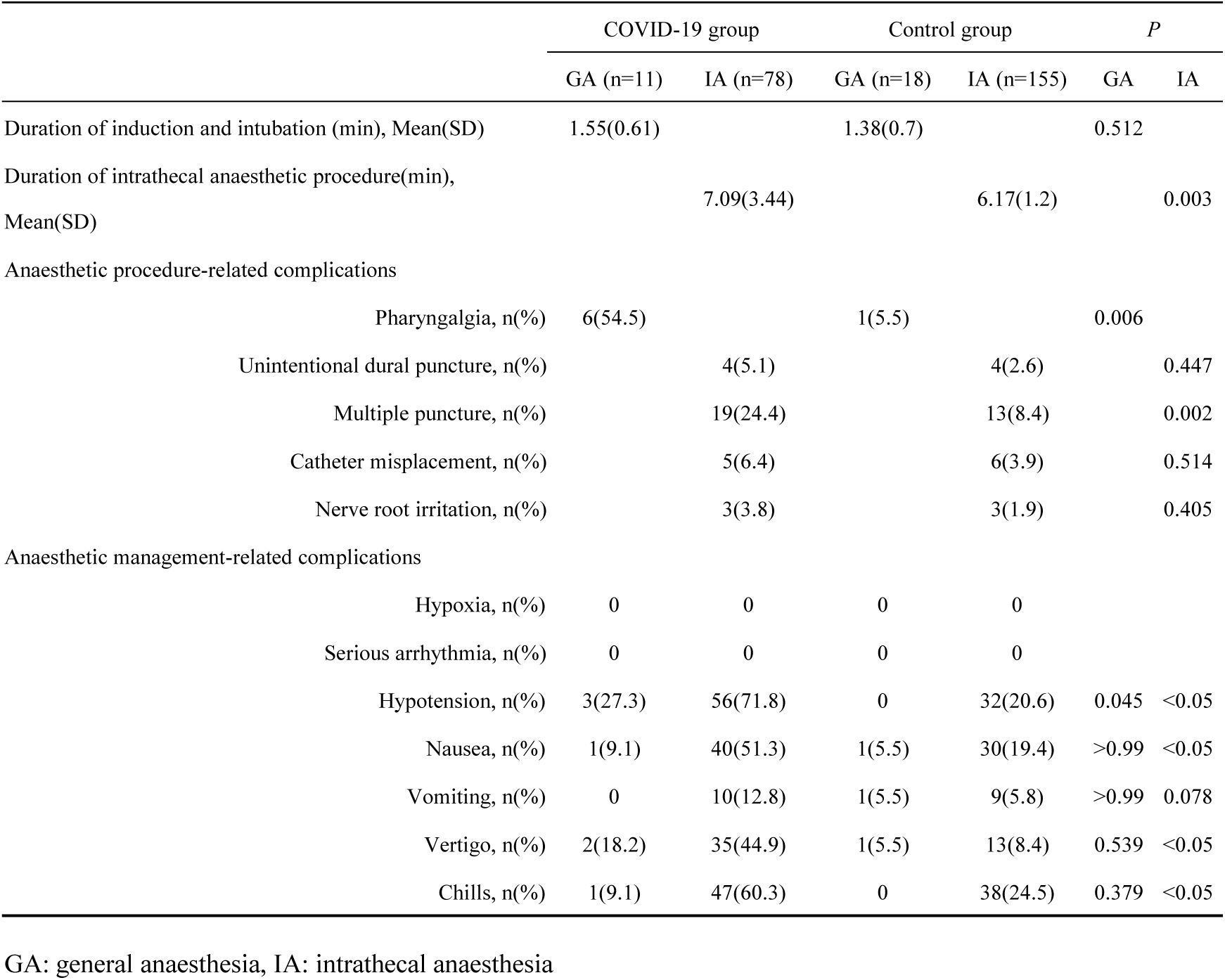
The comparison of complications between two groups after matching

### Maternal outcomes

The length of surgical operation and duration of hospital stay were significantly prolonged in the COVID-19 group compared with the Control group (both *p*<0.05). The intraoperative blood loss in the COVID-19 group was higher than that in the Control group, but there was no difference (*p*>0.05). Besides, the frequency use of intraoperative oxytocin and the time of postoperative oxygen therapy were significant higher in the COVID-19 group than that in the Control group (both *p*<0.05). There were no patients required peripartum admission to the intensive care unit (ICU) and no patients was diagnosed with postpartum depression in both the groups (**Table 5**).

**Table 5.** Maternal clinical outcomes between two groups after matching

|  | COVID-19 group |  | Control group |  | *P* |  |
| --- | --- | --- | --- | --- | --- | --- |
|  | GA (n=11) | IA (n=78) | GA (n=18) | IA (n=155) | GA | IA |
| Length of surgical operation (min), mean(SD) | 69.12(15.40) | 60.19(21.77) | 57.25(14.18) | 55.03(10.3) | 0.044 | 0.015 |
| Duration of hospital stay (days), mean(SD) | 13.82(2.41) | 12.20(3.88) | 7.04(1.66) | 6.58(1.74) | <0.05 | <0.05 |
| Intraoperative blood loss (ml), mean(SD) | 257.18(70.96) | 240.00(65.4) | 231.50(47.49) | 227.31(55.07) | 0.252 | 0.121 |
| Intraoperative utilization of oxytocin, n(%) | 9(81.8) | 33(42.3) | 11(77.8) | 41(26.5) | 0.412 | 0.017 |
| Postoperative oxygen therapy (hours), mean(SD) | 23.80(6.25) | 15.34(4.9) | 10.07(5.22) | 8.91(3.43) | <0.05 | <0.05 |
| Admission to intensive care unit, n(%) | 0 | 0 | 0 | 0 |  |  |
| Postpartum depression, n(%) | 0 | 0 | 0 | 0 |  |  |

GA: general anaesthesia, IA: intrathecal anaesthesia

### Neonatal outcomes

Neonatal, Apgar scores ≤ 8 was defined as low Apgar scores in the present study. For patients underwent general anaesthesia, the rate of low Apgar scores of newborns at 1 and 5 minutes was significantly higher in the COVID-19 group than that in the Control group (both *p*<0.05). However, no significant differences were found in the low Apgar scores rate at 10 minutes between the two groups (all *p*>0.05). As for cases with intrathecal anaesthesia, there was a significant increase in the rate of low Apgar scores for newborns at 1 minutes but not 5 minutes in the COVID-19 group than that in the Control group. All baby Apgar scored were higher than 8 at 10 minutes. Severe neonatal asphyxia and low birth weight (< 2500 g) were not different between the two groups. A total of 9 cases required admission to the NICU in the Control group, and all of 134 cases newborns of COVID-19 parturients were admitted to the NICU for precaution further care. None newborn was with SARS-CoV-2 infection in the COVID-19 groups (**Table 6**), and no neonatal death was in the both groups in present study.

**Table 6.** Neonatal clinical outcomes between two groups after matching

|  | COVID-19 group |  | Control group |  | *P* |  |
| --- | --- | --- | --- | --- | --- | --- |
|  | GA (n=11) | IA (n=79) | GA (n=18) | IA (n=156) | GA | IA |
| Apgar scores ≤ 8 |  |  |  |  |  |  |
| 1 minutes, n(%) | 8(72.7) | 4(5.1) | 2(11.1) | 1(0.6) | <0.05 | 0.045 |
| 5 minutes, n(%) | 5(45.5) | 1(1.3) | 1(5.6) | 1(0.6) | 0.019 | >0.99 |
| 10 minutes, n(%) | 1(9.1) | 0 | 0 | 0 | 0.400 |  |
| Severe neonatal asphyxia, n(%) | 0 | 0 | 0 | 0 |  |  |
| Birth weight (g), mean(SD) | 3180(195) | 3210(182) | 3060(126) | 3250(153) | 0.053 | 0.077 |
| Infant of birth weight <2500g, n(%) | 0 | 0 | 0 | 0 |  |  |
| Admission to the neonatal ICU, n(%) | 11(100) | 79(100) | 5(27.8) | 4(2.6) | <0.05 | <0.05 |
| SARS-CoV-2 positive, n(%) | 0 | 0 | 0 | 0 |  |  |
| Neonatal death, n(%) | 0 | 0 | 0 | 0 |  |  |

GA: general anaesthesia, IA: intrathecal anaesthesia

## Discussion

In this study, we comparatively analyzed the epidemiological data, anaesthetic techniques and management, complications and outcomes of maternal patients with and without COVID-19 in the 3 hospitals in Hubei Province. The results indicated that the high rates of emergency and general anaesthesia use were likely due to the high risk for parturients and newborn.

Consisting with previous reports^9^, the clinical manifestation of parturients with COVID-19 had mild symptoms and none severe phenotype if the disease was in our study. Thus, the intrathecal anaesthesia was the first choice for parturients with COVID-19 in order to reduce medical staff exposing to virus via patient’s airway. Spinal anaesthesia is a safe and quick technique for cesarean delivery due to its quality of sensory blockade and reliability. However, the risk of high level of sensory blockade and the high incidence and degree of hypotension may be the disadvantages when compared with epidural anaesthesia. Although none of the mothers who received intrathecal anaesthesia for delivery had any complications in this study, considering that SARS-CoV-2 virus may also invade the central nervous system^10^, epidural anaesthesia seems to be superior for patients with COVID-19. Due to the limited tactile sensation of PPE, it is necessary to use ultrasound guide epidural procedure for avoiding unintentional dural puncture and subarachnoid catheterization.

In the period of COVID-19 outbreak, routine prenatal examination was limited by traffic control and isolation policy in place, and hence pathological pregnancy can’t be found timely, which may be the reasons for the high emergency caesarean delivery and general anaesthesia use in the COVID-19 patients in this study. Although general anaesthesia would induce or exacerbate pulmonary complications in parturients^11^ with COVID-19, the cesarean delivery under general anaesthesia is necessarily to be used for fetal distress, potential contraindication of intrathecal anaesthesia, as well as intrathecal anaesthesia failure. The rapid-sequence induction by using propofol, rocuronium and remifentanil was used in our COVID-19 patients and the excellent maternal hemodynamic stability was noted. However, the possible risk of short-time neonatal depression and required short periods of mask ventilation or tactile stimulation of the neonate were found. However, in this study, either intravenous or inhalational induction was not difference in term of the safety of maternal and neonatal and the risk infection of medical staff. To avoid hemorrhage resulted from uterine atony, inhalational sevoflurane was switched into intravenous anaesthetics after umbilical cordligation. Notably, PPE increased the difficult in airway management and it was necessary for the use of visual laryngoscopes, and the supraglottic airway was considered as a remedy for failed tracheal intubation in emergency general anaesthesia^12^.

There was a high incidence of intraoperative hypotension in the COVID-19 patients. The risk of hypotension was likely due to anaesthesia induced sympathetic inhibition, preoperative fasting. A recent study showed that the S protein of SARS-CoV-2 binds to the ACE2 receptor^13^, suggesting that the high susceptible of the circulatory system to SARS-CoV-2 infection might be one of the reasons of hypotension in this study. Together with liquid loading and left lateral position, prophylactic phenylephrine was effective and safe in maintaining blood pressure. Prophylactic uterotonic agents (oxytocin) treatment and controlled umbilical cord traction during placenta delivery were also the effective ways to prevent hemorrhage-induced hypotension. As such, it is necessary for COVID-19 parturients to have a contingency plan in place for the high risk of obstetric hemorrhage.

Although a little evidence supported that supplemental oxygen improves maternal and neonatal outcomes^14^, parturients with COVID-19 were routinely treated with oxygen during operation to prevent maternal and infant hypoxia. However, considering the oxygen free radical production and atelectasis, continuous inspiration of high oxygen concentration was not used^15^. It is critical to apply the lung expansion and positive end-expiratory pressure technique for COVID-19 patients receiving general anaesthesia to eliminate atelectasis formation^16^. As for parturients with COVID-19 undergoing intrathecal anaesthesia, the deep-breathing exercises were also encouraged.

Our data showed a down-trend in the number of parturients with COVID-19, which implied the importance of proper home quarantine and personal protection to reduce exposure, and ultimately reduced the chance of COVID-19 infection in pregnant women. Besides, the delivery of infected parturients to designated hospitals was to ensure the safety of the pregnant women and their newborns, while preventing and controlling newborns’ infection with SARS-Cov-2. Potential multi-target organ damage caused by SARS-CoV-2 infection in pregnant women may increase the chance of COVID-19 parturients received caesarean section. Taken more precaution care for those patients are necessary. Cesarean delivery increased the risk of microenvironmental aerosols. Consequently, the fetus should be cleaned of amniotic fluid and blood from the mouth and nose and then transferred to incubator rapidly after delivery to reduce neonatal infection. Additionally, SARS-CoV-2 virus may also invade the central nervous system, indicating that the risk of maternal nervous system injury cannot be ignored. While, none was diagnosed with postpartum depression in the present study, which might be attributed to the implementation of multimode analgesia management in our patient population^17^.

In summary, the current data suggested that the proper home quarantine and personal protection can help to reduce the risk of parturients infected with SARS-Cov-2. Anaesthesia-related complications occurred more frequently in the COVID-19 parturients, while their newborn born also have a high risk of distress and high admission to NICU. From our experience, spinal or epidural anaesthesia can be safely used for such patients. General anaesthesia in general results in more hypotension and open airway for a high risk of virus exposure to medical staff and thus it should be avoided unless it is inevitably used for emergency case.

### Data sharing

No additional data available.

The manuscript‘s guarantor affirms that the manuscript is an honest, accurate, and transparent account of the study being reported; that no important aspects of the study have been omitted; and that any discrepancies from the study as planned and registered have been explained.

## Data Availability

All data generated or analyzed during this study are included in this article. No additional data available.

